# Human genetic evidence to inform clinical development of interleukin-6 signaling inhibition for abdominal aortic aneurysm

**DOI:** 10.1101/2024.09.14.24313670

**Authors:** Stephen Burgess, Héléne T. Cronjé, Emil deGoma, Yung Chyung, Dipender Gill

## Abstract

**Background:** Abdominal aortic aneurysm (AAA) represents a significant cause of mortality, yet no medical therapies have proven efficacious. The aim of the current study was to leverage human genetic evidence to inform clinical development of interleukin-6 (IL6) signaling inhibition for treatment of AAA.

**Methods:** We focused on rs2228145, a missense variant in the *IL6R* gene region whose associations are expressed per additional copy of the C allele, corresponding to the genetically-predicted effect of IL6 signaling inhibition. We consider genetic associations with AAA risk in the AAAgen consortium (39,221 cases, 1,086,107 controls) and UK Biobank (2215 cases, 365,428 controls). To validate against known effects of IL6 signaling inhibition, we present associations with rheumatoid arthritis, polymyalgia rheumatica, and severe COVID-19. To explore mechanism specificity, we present associations with thoracic aortic aneurysm, intracranial aneurysm, and coronary artery disease. We further evaluated associations with measures of the abdominal aorta in UK Biobank, and explored genetic associations in clinically-relevant subgroups of the population.

**Results:** We observed strong genetic associations with AAA risk in the AAAgen consortium and in UK Biobank: odds ratio (OR) 0.91 (95% confidence interval [CI]: 0.90 to 0.92, p = 4×10^-30^) and OR 0.90 (95% CI: 0.84, 0.96, p=0.0007), respectively. The association with AAA risk in UK Biobank was linear in the number of minor alleles: OR 0.91 (95% CI: 0.83, 1.00) in heterozygotes and OR 0.80 (95% CI: 0.71, 0.92) in minor homozygotes. The association was similar for fatal AAA, but with greater uncertainty due to the lower number of events. The association with AAA was of greater magnitude than associations with coronary artery disease and even rheumatologic disorders for which IL6 inhibitors have been approved. No strong associations were observed with thoracic aortic aneurysm, intracranial aneurysm, or abdominal aorta diameter in the general population without AAA. Associations attenuated towards the null in populations with concomitant inflammatory or connective tissue disease.

**Conclusions:** This drug target Mendelian randomization study supports that IL6 signaling inhibition will be efficacious for treating AAA, but not other types of aneurysmal disease. These findings serve to help inform clinical development of IL6 signaling inhibition for AAA treatment.

## Introduction

Abdominal aortic aneurysm (AAA) is defined as a permanent localized dilatation of the abdominal aorta to more than 3cm and has a prevalence of approximately 5% in individuals aged more than 60 years.^1^ Mortality from ruptured AAA is estimated to be approximately 90%,^2,3^ and other than for modifying cardiovascular risk factors, no efficacious pharmacotherapies are available for AAA treatment.^4^ The mainstay of intervention is therefore surgical repair, despite which global AAA-related mortality remains to be at approximately 150,000 to 200,000 deaths per year.^5,6^

Drug development can be slow, expensive, and inefficient,^7^ with the high failure rate largely attributable to insufficient efficacy or unacceptable safety profiles.^8^ As the majority of drug targets are proteins, which are coded for by genes, human genetic data offer the opportunity to dramatically improve the probability of successful drug development.^9^ At a population level, naturally occurring genetic variation in the gene coding for a drug target protein can be used to study the effect of pharmacologically perturbing that protein.^10^ This drug target Mendelian randomization paradigm can be used to inform various aspects of clinical development,^11^ including efficacy, secondary indications, adverse effects, effect heterogeneity, and biomarkers of target engagement.^12^ In this way, drug target Mendelian randomization has been used to inform on the effects of various cardiovascular disease (CVD) drug targets, including for lipid-lowering, antihypertensive, anti-diabetic, and anticoagulant agents. ^11^

The drug target Mendelian randomization paradigm has been used extensively to study the effect of interleukin-6 (IL6) signaling inhibition on CVD outcomes.^13–15^ The genetic evidence supporting its efficacy has contributed to the pursuit of IL6 signaling inhibition as a therapeutic target in CVD, with one phase 2 study (NCT06362759) and five phase 3 clinical studies currently ongoing (NCT05021835, NCT06118281, NCT05636176, NCT05485961, NCT06200207).^16^ However, while there is also genetic support for the efficacy of IL6 signaling inhibition in AAA,^17^ this has remained limited in nature with key questions yet unanswered. The aim of this work was therefore to leverage the drug target Mendelian randomization paradigm to inform clinical development of IL6 signaling inhibition for AAA. Specifically, we aimed to affirm the genetic evidence for efficacy of IL6 signaling inhibition in AAA, compare against established positive control outcomes and coronary artery disease (CAD), investigate specificity for different types of aneurysmal disease and effects on abdominal aorta diameter more generally, and explore heterogeneity of effect across population subgroups. These findings collectively serve to inform clinical development efforts of IL6 signaling inhibition for the treatment of AAA.

## Methods

### Overview

As the instrument for IL6 signaling inhibition, we leverage a genetic variant in the *IL6R* gene region that has previously been shown to mimic pharmacological IL6 signaling inhibition.^14^ To investigate potential clinical effects of inhibiting IL6 signaling, we present associations of this variant with traits and diseases from a variety of data sources, including publicly-available summarized datasets and individual-level data from UK Biobank. The summarized datasets are larger and have more cases, resulting in more precise estimates. The individual-level data analyses allow the exploration of more specific outcomes, such as fatal abdominal aortic aneurysm, and subgroup analyses in specific strata of the population. Additionally, a subset of UK Biobank participants have undergone magnetic resonance imaging (MRI), enabling investigation of genetic associations with measures of the abdominal aorta. An overview of all analyses and datasets is provided as **Figure 1**.

**Figure 1.**
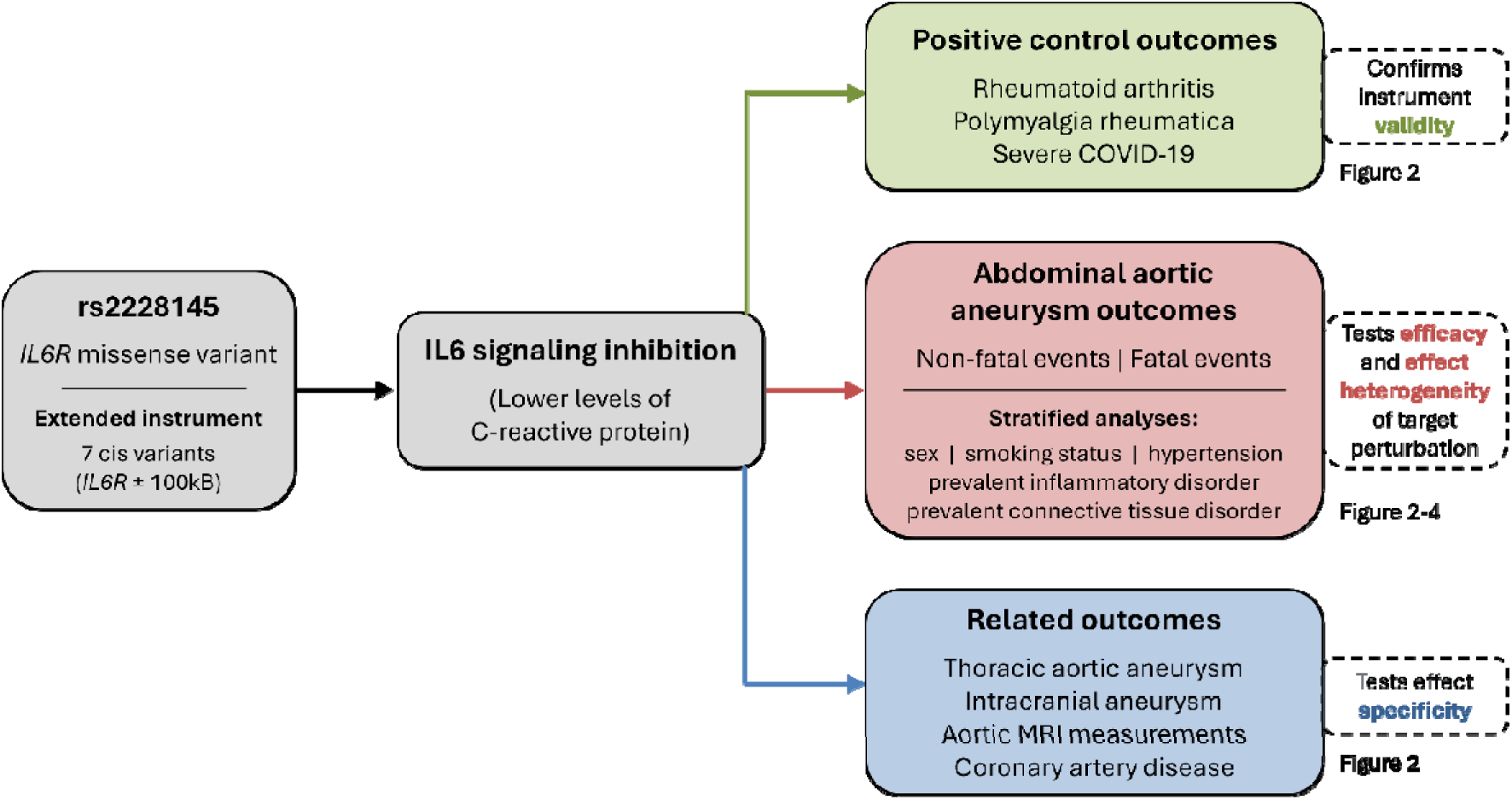
Overview of analyses. The Mendelian randomization framework relies on the following core instrumental variable assumptions: 1) the genetic instrument strongly relates to the exposure; 2) there are no confounding pathways linking variant and outcome, and; 3) the genetic instrument affects the outcome only through the exposure and not through independent pathways. IL6: interleukin-6, IL6R: interleukin-6 receptor.

### Genetic instrument

We focused on rs2228145 (previously also called rs8192284), a missense variant located at chr1:154426970 on GRCh37, chr1:154454494 on GRCh38 with minor allele frequency of 42% in the European ancestry UK Biobank analytic sample considered here. It is the lead variant associated with IL6 levels in the *IL6R* gene region (**Supplementary** Figure 1). Associations are given per additional copy of the C allele, which is the minor allele and is associated with lower levels of C-reactive protein (0.093 units lower log-transformed C-reactive protein, p < 10^-400^).^18^ Hence estimates correspond to the genetically-predicted effect of increasing interleukin-6 signaling inhibition. We also considered estimates based on an extended instrument consisting of 7 variants in the *IL6R* gene region and its neighborhood (±100 kilobase pairs) previously demonstrated to be conditionally associated with C-reactive protein levels (**Supplementary Table 1**).^14^

### Summarized datasets

The primary outcome was AAA. Associations were obtained from a meta-analysis of 17 individual GWAS by the AAAgen Consortium including 39,221 cases and 1,086,107 controls.^19^ We performed colocalization between IL6 and AAA risk using the coloc method,^20^ taking genetic associations with IL6 from the SCALLOP consortium (N=14,242)^21^ and using the default priors from the coloc package.^22^ Colocalization is a statistical approach to assess whether the genetic predictors of two traits overlap (known as colocalization) or are distinct (known as non-colocalization). The coloc method reports two key outputs: the posterior probability of colocalization (PP-H4) and the posterior probability of non-colocalization (PP-H3).

High values (close to 1) of PP-H4 indicate colocalization, which is supportive of a causal relationship; high values of PP-H3 indicate non-colocalization, which opposes a causal relationship; low values of both PP-H3 and PP-H4 indicate lack of strong evidence supporting or opposing a causal relationship.^23^

As positive controls, we present associations with rheumatoid arthritis (GWAS consortium, 35,871 cases, 240,149 controls;^24^ FinnGen, 15,223 cases, 138,246 controls),^25^ polymyalgia rheumatica (UK Biobank, 2,460 cases, 433,511 controls),^26^ and severe COVID-19 (COVID-19 Host Genome Initiative round 7, 18,152 cases, 1,145,546 controls),^27^ as IL6 signaling inhibition is known from randomized trials to offer therapeutic benefit in these diseases.^28–30^

For comparison with other aneurysmal and CVDs, we present associations with thoracic aortic aneurysm (Michigan Genomics Initiative, 1,351 cases, 18,295 controls),^31^ intracranial aneurysm (GWAS Consortium, 10,754 cases, 306,882 controls)^32^, and CAD (CARDIoGRAMplusC4D, 210,842 cases, 1,167,328 controls).^33^ We note that all summarized datasets include UK Biobank participants, with the exception of FinnGen and the Michigan Genomics Initiative.

### Individual-level data in UK Biobank

The UK Biobank cohort comprises around 500,000 participants (94% of self-reported European ancestry) aged 40 to 69 years at baseline.^34^ They were recruited between 2006-2010 in 22 assessment centers throughout the UK, and followed up until November 2022 or their date of death. We performed detailed quality control procedures on UK Biobank participants and on genetic variants as described previously,^35^ restricting analyses to unrelated participants (that is, more distant than third degree relatives) of European ancestries.

Abdominal aortic aneurysm in UK Biobank was defined as having an International Statistical Classification of Diseases and Related Health Problems (ICD)-10 code of I71.3 or I71.4 (AAA, either ruptured or without mention of rupture) or I71.5 or I71.6 (thoracoabdominal aortic aneurysm, either ruptured or without mention of rupture), or the equivalent ICD-9 code (441.3, 441.4, 441.5, 441.6) in their hospital episode statistics or death certificate. The secondary outcome was fatal AAA, defined as having one of the relevant ICD codes in their death certificate. We also performed sensitivity analyses excluding thoracoabdominal aortic aneurysms (252 events excluded).

Amongst MRI measurements, we assessed associations with derived variables “abdominal descending aorta (horizontal diameter)” (UK Biobank field ID 31090) and “abdominal descending aorta (vertical diameter)” (field ID 31091).^36^ These correspond to the transverse and anteroposterior diameters of the abdominal aorta respectively, as measured by axial MRI.

### Stratification variables

We estimated associations with AAA in UK Biobank in subgroups of the population, stratifying by five separate variables: sex (male vs female), hypertension (defined as systolic blood pressure > 140 mmHg and diastolic blood pressure > 90 mmHg at baseline, or usage of hypertensive medication at baseline), smoking status (ever regular smoker vs never regular smokers), any rheumatologic disorder (present vs absent), and any connective tissue disorder (present versus absent).

A never regular smoker was defined as answering the touchscreen question “In the past, have you ever smoked tobacco?” with the response “I have never smoked” or “Just tried once or twice” as opposed to “smoked occasionally” or “smoked on most or all days” at baseline, and never contradicting this answer at a future survey.

Any rheumatologic disorder was defined as ankylosing spondylitis, anti-neutrophil cytoplasmic antibody-associated vasculitis, Behçet disease, Cogan’s syndrome, giant cell arthritis, relapsing polychondritis, rheumatoid arthritis, sarcoidosis, systemic lupus erythematosus, or Takayasu arteritis, defined using ICD codes and self-reported information (**Supplementary Table 2**).

Any connective tissue disorder was defined as Marfan syndrome, Ehlers-Danlos syndrome, or any non-specific connective tissue disorder, defined using ICD codes and self-reported information (**Supplementary Table 2**).

### Statistical analyses

All associations in UK Biobank were obtained by logistic regression (for disease outcomes) or linear regression (for MRI measurements) adjusted for age, sex, and 10 genomic principal components of ancestry using an additive genetic model. We repeated the primary analysis using a factorial model regarding major homozygotes as the reference group, and providing separate estimates for heterozygotes and minor homozygotes. Analyses for the primary instrument are reported as genetic associations per additional C-reactive protein decreasing allele. Analyses for the extended instrument were performed using exposure data on genetic associations with C-reactive protein,^18^ and implemented using the inverse-weighted method accounting for correlation between variants, and a fixed-effect model. The genetic correlation matrix was estimated in the analytic sample of UK Biobank participants. All analyses were performed in R (version 4.3.3). All p-values are two-sided.

### Ethical approval and participant consent

Individual-level analyses on data from the UK Biobank were approved by its Research Ethics Committee and Human Tissue Authority research tissue bank under application number 7493. For analysis of prior published genetic associations, summary statistics were gathered from studies that had obtained appropriate independent ethical approval and participant consent for analyses and distribution of summary-level data, as described in the original publications.

### Data and code availability

Summary statistics used in our analyses can be accessed through the citations provided. Requests to access UK Biobank data can be made by bona fide researchers from any sector. More information can be found at https://www.ukbiobank.ac.uk/enable-your-research. Statistical code for the analyses undertaken in this work are available from the corresponding author upon reasonable request. This study is reported using the Strengthening the Reporting of Observational Studies in Epidemiology-MR guidelines (**Checklist**).^37^

## Results

### Demographic information on UK Biobank analytic sample

A total of 367,643 unrelated UK Biobank participants of European ancestries were included in individual-level data analyses. In total, 2,215 individuals had an AAA diagnosis, and 143 had a fatal AAA diagnosis. A total of 198,838 individuals were female (54.1%), 120,051 individuals had hypertension (32.6%), 169,788 individuals were ever-smokers (46.2%), 11,551 had an rheumatologic disorder (3.1%), and 505 had a connective tissue disorder (0.1%). Demographic information on participants is provided in **Table 1**.

**Table 1.** Demographic information on UK Biobank participants in the analytic sample divided by stratification variables.

| Trait | Overall | Sex |  | Smoking status |  | Hypertension |  | IDs | CTDs |
| --- | --- | --- | --- | --- | --- | --- | --- | --- | --- |
|  |  | Male | Female | Ever | Never | Yes | No |  |  |
| **Sample size (N)** | 367,643 | 168,748 | 198,838 | 169,788 | 197,853 | 120,051 | 244,971 | 11,551 | 505 |
| **Age (years)** | 57.2 | 57.4 | 57.0 | 58.0 | 56.5 | 59.9 | 55.9 | 59.6 | 56.1 |
| **Sex (% female)** | 54.1 | 0 | 100 | 48.6 | 58.8 | 44.2 | 59.1 | 62.2 | 68.9 |
| **AAA events (N)** | 2,215 | 1,865 | 350 | 1815 | 400 | 1,377 | 822 | 137 | 13 |
| **Fatal AAA (N)** | 143 | 126 | 17 | 129 | 14 | 93 | 50 | 9 | 0 |
| **BMI (kg/m<sup>2</sup>)** | 27.4 | 27.8 | 27.0 | 27.7 | 27.1 | 29.2 | 26.4 | 28.3 | 26.7 |
| **LDL-c (mmol/L)** | 3.57 | 3.49 | 3.64 | 3.54 | 3.59 | 3.45 | 3.62 | 3.52 | 3.51 |
| **ApoB (g/L)** | 1.03 | 1.03 | 1.04 | 1.03 | 1.03 | 1.02 | 1.04 | 1.03 | 1.02 |
| **HbA1c (mmol/mol)** | 35.9 | 36.3 | 35.6 | 36.4 | 35.5 | 37.6 | 35.1 | 36.6 | 35.4 |

AAA: abdominal aortic aneurysm, ApoB: apolipoprotein-B, BMI: body mass index, CTD: connective tissue disorders, HbA1c: glycated hemoglobin, IDs: rheumatologic disorders, LDL-c: low-density lipoprotein cholesterol.

While around 30,000 UK Biobank participants have undergone MRI scanning, data on aortic MRI were only available for 619 UK Biobank participants, none of whom had an AAA diagnosis. Compared to the overall UK Biobank population, these participants were slightly younger (average age 53.5 versus 57.2 years) with a similar sex balance (female 53.2% versus 54.1%), and had similar average levels of cardiovascular risk factors (**Supplementary Table 3**).

### Genetic evidence for efficacy of IL6 signaling inhibition in AAA

The genetic association with AAA in the AAAgen consortium was an odds ratio (OR) of 0.91 (95% confidence interval [CI]: 0.90 to 0.92, p = 4×10^-30^) per additional copy of the C allele of rs2228145 (**Figure 2**). The association in UK Biobank was similar: 0.90 (95% CI: 0.84, 0.96, p=0.0007). The genetic association with fatal AAA risk in UK Biobank was similar, but had wider confidence intervals due to the lower number of events: 0.87 (95% CI: 0.69, 1.11, p=0.26). Associations in UK Biobank were similar in a sensitivity analysis excluding thoracoabdominal aortic aneurysm cases: 0.90 (95% CI: 0.84, 0.96, p=0.001). Associations were approximately additive considering genetic subgroups separately in a factorial model, with an OR of 0.91 (95% CI: 0.83, 1.00, p = 0.047) for heterozygotes and 0.80 (95% CI: 0.71, 0.92, p = 0.0009) for minor homozygotes (**Figure 3)**.

**Figure 2.**
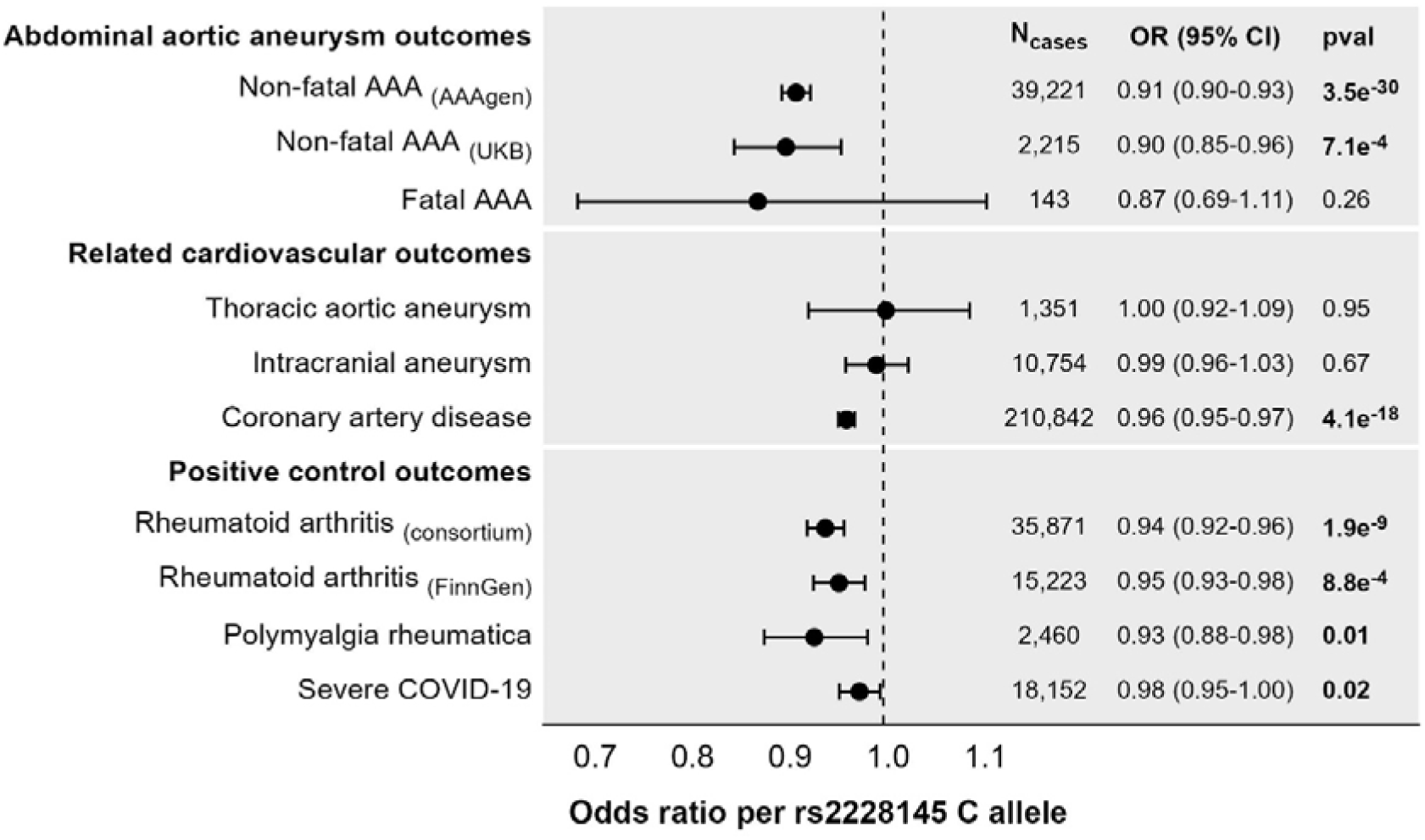
Genetic associations with outcomes estimated in summarized and individual-level data. Estimates represent odds ratio (OR) per additional copy of the C allele for rs2228145, corresponding to the genetically-predicted effect of increasing interleukin-6 signalling inhibition. Positive control outcomes are outcomes for which IL6 inhibitors have proven efficacious. CI: confidence interval; pval: p-value.

**Figure 3.**
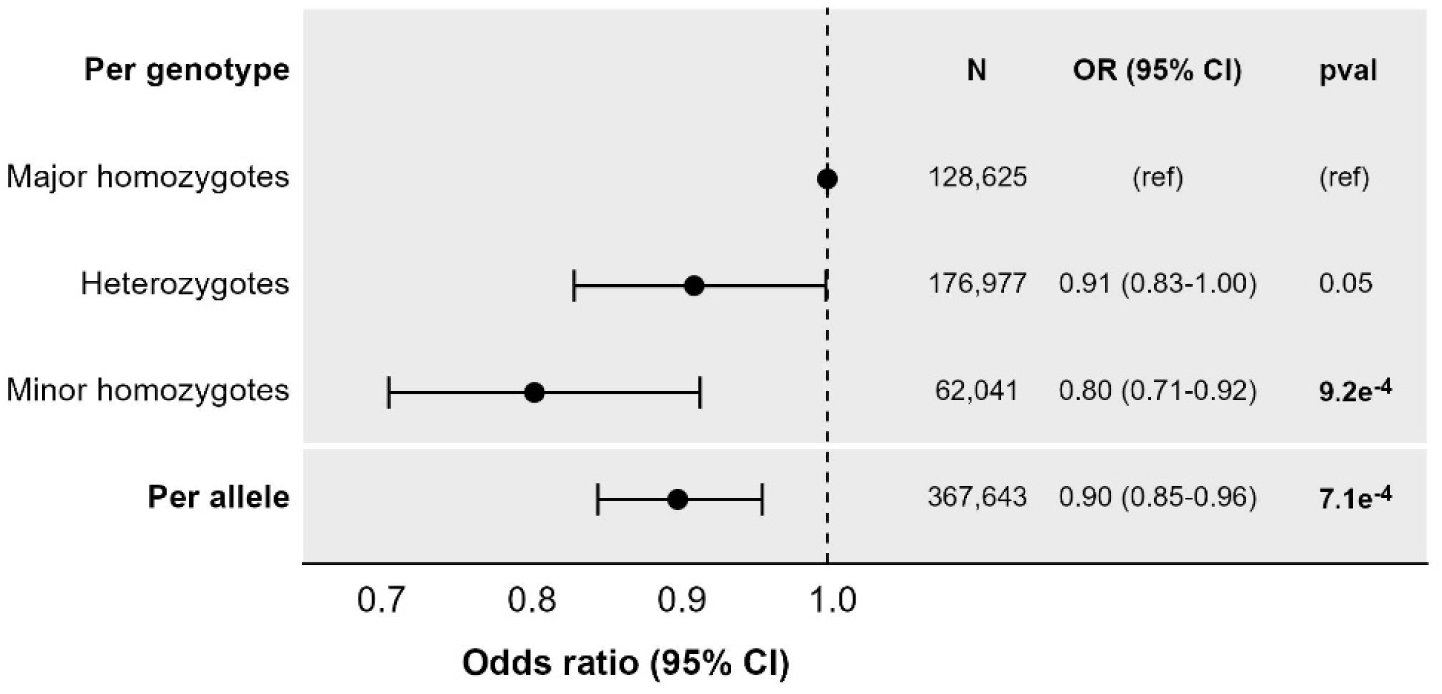
Genetic associations with abdominal aortic aneurysm in UK Biobank from per allele and factor models. Estimates represent odds ratio of an abdominal aortic aneurysm per additional copy of the C allele for rs2228145 (per allele model), or for heterozygotes (AC genotype) and minor allele homozygotes (CC genotype) compared with major allele homozygotes (AA genotype). CI: confidence interval; pval: p-value.

The estimate using the extended instrument of 7 variants in the *IL6R* gene region was OR of 0.93 (95% CI: 0.91, 0.94) per 0.1 unit lower log-transformed C-reactive protein. All 7 variants provided supportive evidence of a causal effect with the exception of rs12083537, which was an outlier, and hence may be pleiotropic in terms of its genetic association with the outcome (**Supplementary** Figure 2). On the same scale, the estimate using the rs2228145 variant only was 0.90 (95% CI: 0.89, 0.92). Given limited benefit in terms of precision, and the possibilities of including pleiotropic variants and overfitting using the extended instrument,^38^ we performed all further analyses using the rs2228145 variant only.

There was strong evidence of colocalization for the genetic association with IL6 levels and AAA risk at the *IL6R* gene locus, supportive of a causal relationship: posterior probability of colocalization (PP-H4) = 0.996.

### Genetic evidence for efficacy of IL6 signaling inhibition on positive control outcomes

The genetic association with rheumatoid arthritis was 0.94 (95% CI: 0.92, 0.96, p = 2×10^-9^) in the GWAS consortium, and 0.95 (95% CI: 0.93, 0.98, p = 0.0009) in FinnGen. The association with polymyalgia rheumatica was 0.93 (95% CI: 0.88, 0.98, p = 0.012). The association with severe COVID-19 was 0.98 (95% CI: 0.95, 1.00, p = 0.023). Associations with these positive controls provide evidence that the genetic associations are a reliable guide for the impact of IL6 signaling inhibition in clinical trials.

### Genetic evidence for efficacy of IL6 signaling inhibition on other aneurysmal and cardiovascular outcomes

Associations with other aneurysmal diseases were 1.00 (95% CI: 0.92, 1.09, p = 0.95) for thoracic aortic aneurysm, and 0.99 (95% CI: 0.96, 1.03, p = 0.67) for intracranial aneurysm. It appears that genetic evidence for benefit of IL6 signaling inhibition on aneurysm risk is specific to AAA.

The association with CAD was 0.96 (95% CI: 0.95, 0.97, p = 4×10^-18^) for CAD. We note that associations with AAA are at least twice as strong as CAD.

Among UK Biobank participants with available MRI data and without AAA, mean horizontal diameter of the abdominal descending aorta was 21.7 mm (interquartile range 20.0 to 23.3) and mean vertical diameter was 22.1 mm (interquartile range 20.4 to 23.7). There was no significant association between IL6 signaling inhibition and the diameter of the abdominal aorta assessed by MRI. Genetic associations with aortic diameter available in the general UK Biobank population were 0.13 mm (95% CI: –0.11, 0.36, p = 0.29) for abdominal descending aorta (horizontal diameter), and 0.22 mm (95% CI: –0.01, 0.45, p = 0.06) for abdominal descending aorta (vertical diameter). This represents the absence of genetic evidence implicating IL6 signaling inhibition in abdominal aorta dimensions outside the setting of AAA, although with the caveat of limited statistical power.

### Stratified analyses in UK Biobank

Stratified analyses were performed in UK Biobank participants. The genetic association with abdominal aortic aneurysm risk was slightly stronger in males: 0.89 (95% CI: 0.84, 0.96, p=0.0009) than in females: 0.93 (95% CI: 0.80, 1.09, p=0.37), although there was no statistical evidence for a difference between estimates (p=0.61). Similar findings were obtained regardless of hypertension or smoking status. Among individuals with inflammatory or connective diseases, estimates were attenuated towards the null although the confidence intervals still overlapped (**Figure 4**).

**Figure 4.**
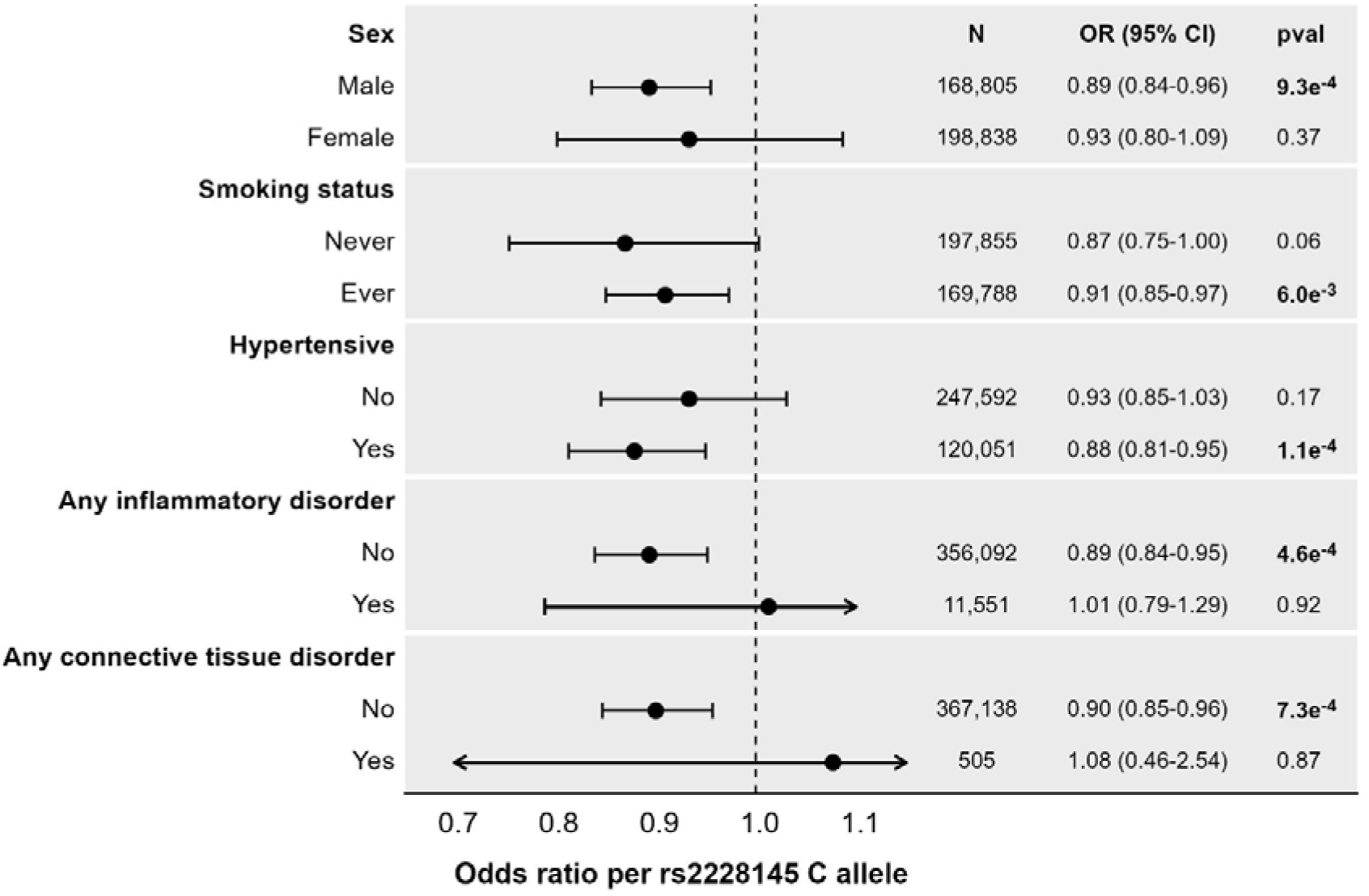
Stratified genetic association estimates with abdominal aortic aneurysm in UK Biobank. Estimates represent odds ratio (OR) of an abdominal aortic aneurysm event per additional copy of the C allele for rs2228145, corresponding to the genetically-predicted effect of greater interleukin-6 signalling inhibition. CI: confidence interval; pval: p-value.

## Discussion

We used a genetic variant mimicking the effects of IL6 signaling inhibition in the Mendelian randomization paradigm and identified evidence supporting protective effects on AAA risk. These findings are consistent with previous work,^14,17^ but make a number of important advances. Firstly, we demonstrate an additive effect for each additional rs2228145 C allele (mimicking IL6 signaling inhibition) on AAA risk reduction, consistent with IL6 signaling driving AAA pathophysiology. Secondly, we show that the magnitude of the Mendelian randomization estimate is similar for risk of fatal AAA as it is for risk of any AAA, supporting similar protective effects on risk of AAA rupture. Thirdly, we find that the Mendelian randomization association was similar in population subgroups stratified by sex, blood pressure, and smoking status, but was attenuated in individuals with AAA related to inflammatory or connective tissue disease, albeit with wider confidence intervals in the diseased subgroups. This supports that protective effects of IL6 signaling inhibition may be specific to AAA arising secondary to atherosclerotic risk factors rather than due to pre-existing inflammatory disease or connective tissue disease. Of note, the Mendelian randomization association for IL6 signaling inhibition with AAA was greater in magnitude than for CAD, suggesting that the beneficial effect in AAA may not be entirely attributable to reduced atherosclerosis. Fourthly, we show that the genetic evidence of effect was specific to AAA and not other types of aneurysmal disease, such as intracranial aneurysm or thoracic aortic aneurysm. Fifthly, we found no strong association of genetically predicted IL6 signaling inhibition with abdominal aortic diameter in individuals without AAA from the general population, again supporting that its effects may be specific to AAA.

Collectively, these findings may be directly employed to inform the clinical development of IL6 signaling inhibition for the treatment of AAA. Previous work has employed Mendelian randomization to study potential biomarkers and adverse effects of inhibiting IL6 signaling.^39^ Clinical trials investigating IL6 signaling inhibition for the treatment of CVD are already underway^16^ and the insights generated in our current study may be used to inform similar endeavors for AAA. Other than addressing general CVD risk factors such as hypertension, dyslipidemia or diabetes mellitus, there are currently no approved pharmacological therapies for the treatment of AAA. The current standard of care is based on monitoring for expansion, with the option of surgical intervention should certain size thresholds be crossed or in the case of rupture, which itself is associated with an approximately 90% mortality rate.^2^ Thus, the availability of efficacious pharmacological therapies for AAA treatment would represent a notable advance in patient care.

There is already a plethora of data implicating IL6 signaling in AAA pathophysiology. Inhibition of IL6 signaling has been shown to limit progression of AAA in animal models,^40^ and is also associated with improved survival.^41^ In humans, IL6 is abundantly expressed in AAA tissue,^42^ and may even be a source of systemic IL6.^43^ Previous Mendelian randomization analyses have supported IL6 signaling in AAA risk,^14,41^ as well as potential effects of IL6 signaling on reducing progression on AAA, although this latter work was limited by low statistical power. Inflammation is a key driver in AAA occurring outside the background of a rheumatological or connective tissue disease,^44^ with inflammatory cell infiltrates observed in the aneurysm wall,^45^ and aneurysm mural thrombus.^46^ It therefore follows that inhibition of IL6 signaling might reduce AAA risk, progression, and rupture.

This work has a number of strengths. Using the Mendelian randomization paradigm, we were able to efficiently generate causal evidence in humans to inform clinical development efforts supporting IL6 signaling inhibition for the treatment of AAA. Specifically, the insights generated here may be used to prioritize the specific type of aneurysmal disease and the target population. To ensure the robustness of our approach, we validated the method with established positive control outcomes where IL6 signaling inhibition has proven efficacious, including for rheumatoid arthritis, polymyalgia rheumatica, and COVID-19. Statistically, we showed that using a biologically validated missense variant as the instrument in Mendelian randomization produced similar estimates to a polygenic *cis*-instrument, and further through colocalization we generated support that genetic confounding through a variant in linkage disequilibrium was unlikely to be explaining the observed associations.

There are also limitations. The Mendelian randomization paradigm employed in this work considers the cumulative lifetime effect of genetic variation on risk of clinical outcomes in a select population. Caution should therefore be taken when extrapolating these findings to assume the effect of a clinical intervention having a larger effect at a discrete timepoint in life in an entirely different population. In this regard, these analyses were largely limited to European genetic ancestry populations, although previous work supports that similar associations may hold in other genetic ancestry populations.^47^ Furthermore, these genetic analyses evaluated the risk of developing AAA but do not directly assess the clinical impact of reduced IL-6 signaling after AAA has already manifested, the clinical setting in which AAA therapies would actually be applied. Finally, a fundamental limitation of all Mendelian randomization analyses is that it assumes any genetic associations between the instrument and outcome are only occurring through the exposure and not some pleiotropic pathway, which can never be proven. It therefore remains possible that our findings may be biased by such pleiotropic effects.

In conclusion, this Mendelian randomization study finds human causal evidence to support the clinical development of IL6 signaling inhibition for the treatment of AAA, including the specific disease subtypes and target populations to be prioritized. Five phase 3 clinical trials of IL6 signaling inhibition for CVD are already underway,^16^ and the weight of the supportive evidence for AAA coupled with the unmet need for efficacious medical therapies signposts this as a promising opportunity for clinical investigation.

## Supporting information

STROBE-MR Checklist

## Data Availability

Summary statistics used in our analyses can be accessed through the citations provided. Requests to access UK Biobank data can be made by bona fide researchers from any sector. More information can be found at https://www.ukbiobank.ac.uk/enable-your-research. Statistical code for the analyses undertaken in this work are available from the corresponding author upon reasonable request.

## Acknowledgements

This research has been conducted using the UK Biobank Resource under Application Number 7439. The authors acknowledge participants and investigators of the UK Biobank and the other cohort studies incorporated in our work. We thank the cohorts and consortia that made their summary-level data publicly available. Data sources are cited throughout this manuscript. Downloads were performed through the following repositories: IL6, https://www.ebi.ac.uk/gwas/studies/GCST90012005; CRP, https://www.ebi.ac.uk/gwas/studies/GCST90029070; AAA, https://csg.sph.umich.edu/willer/public/AAAgen2023/; Rheumatoid arthritis (consortium), https://www.ebi.ac.uk/gwas/studies/GCST90132222; Rheumatoid arthritis (FinnGen), https://www.finngen.fi/en/access_results; Polymyalgia rheumatica, https://www.ebi.ac.uk/gwas/studies/GCST90129454; Severe COVID-19, https://www.covid19hg.org/results/r7/; Thoracic aortic aneurysm, https://www.ebi.ac.uk/gwas/studies/GCST90027266; Intracranial aneurysm, https://cd.hugeamp.org/downloads.html; Coronary artery disease, https://www.ebi.ac.uk/gwas/studies/GCST90132315.

