## Supplementary Materials for "Human genetic evidence to inform clinical development of interleukin-6 signaling inhibition for abdominal aortic aneurysm"

**Supplementary Table 1.** Genetic variants included in the extended instrument.

| rsid | Chromosome and position (GRCh37) |
| --- | --- |
| rs73026617 | chr1:154369981 |
| rs12083537 | chr1:154381103 |
| rs4556348 | chr1:154394296 |
| rs2228145 | chr1:154426970 |
| rs11264224 | chr1:154568086 |
| rs12059682 | chr1:154579585 |
| rs34693607 | chr1:154661369 |

**Supplementary Table 2.** List of codes used to define any rheumatological disorder and any connective tissue disorder.

|  | Codes used | | |
| --- | --- | --- | --- |
| Disorder | **ICD-9** | **ICD-10** | **Self-reported** |
| Rheumatologic disorders | | | |
| Giant cell arthritis | 446.5 | M31.[5,6] | 1376 |
| Takayasu arteritis | 446.7 | M31.4 | - |
| Behçet disease | 136.1 | M35.2 | - |
| Cogan's syndrome | 370.52 | H16.3 | - |
| Anti-neutrophil cytoplasmic antibody-associated vasculitis | 446.4 | M31.3 | 1378 |
| Ankylosing spondylitis | 720.0 | M45.X | 1313 |
| Relapsing polychondritis |  | M94.1 |  |
| Sarcoidosis | 135.X | D86.X | 1371 |
| Systemic lupus erythematosus | 710.0 | M32.[0,1,8,9] | 1424 |
| Rheumatoid arthritis | 714.[0,1,2,81] | M05.[0-3,8,9], M06.[0,8,9] | 1464 |
| Other non-specific rheumatological disorder codes | 279.49 | D89.9 | - |
| Connective tissue disorders | | | |
| Marfan syndrome | 759.82 | Q87.4 | - |
| Ehlers-Danlos syndrome | 757.83 | Q79.6 | - |
| Other non-specific connective tissue disorder codes | - | L94.X | 1373 |

**Supplementary Table 3.** Demographic information on UK Biobank participants in the analytic sample overall compared to those with heart magnetic resonance imaging (MRI) data.

|  | Overall | Heart MRI data |
| --- | --- | --- |
| Sample size (N) | **367,643** | **619** |
| Age (years) | 57.2 | 53.5 |
| Sex (% female) | 54.1 | 53.2 |
| AAA events | 2215 | 0 |
| Fatal AAA events | 143 | 0 |
| Body mass index (kg/m^2^) | 27.4 | 24.4 |
| LDL-cholesterol (mmol/L) | 3.569 | 3.565 |
| Apolipoprotein B (g/L) | 1.034 | 1.013 |
| HbA1c (mmol/mol) | 35.9 | 33.8 |

**Supplementary Figure 1.** Genetic associations with circulating interleukin-6 and C-reactive protein levels in the neighborhood of the *IL6R* gene region (±100kb).

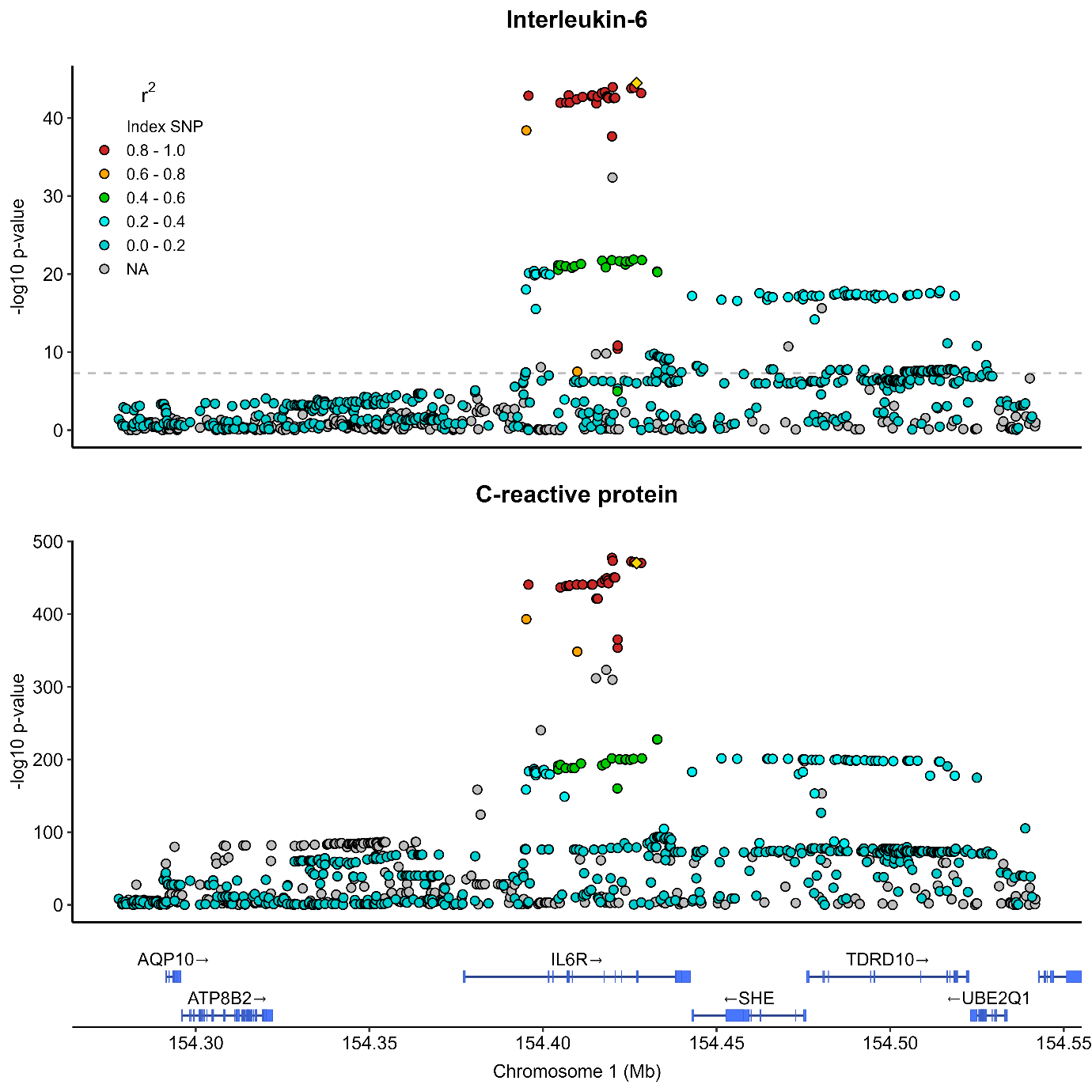

**Supplementary Figure 2.** Scatterplot showing associations of genetic variants in the *IL6R* gene region with C-reactive protein and abdominal aortic aneurysm (AAA) risk

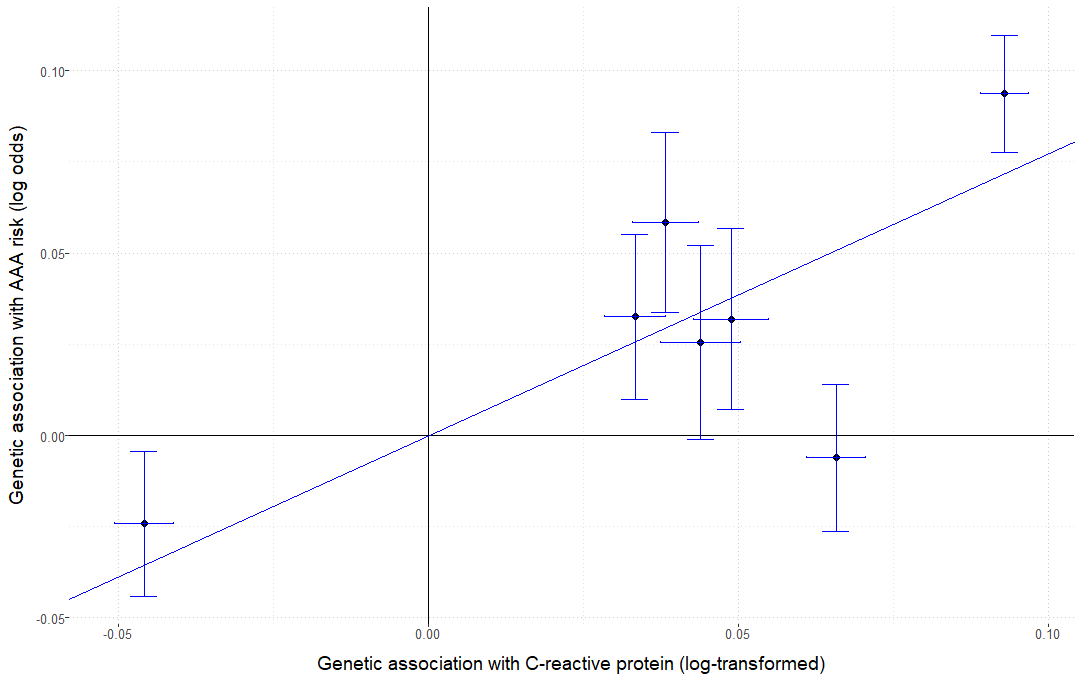
